# Impact of Catheter Ablation on Ventricular Tachycardia Recurrence in Patients with Chagas Cardiomyopathy Following implantable cardioverter-defibrillator Implantation: A Competing Risks Analysis

**DOI:** 10.1101/2025.05.30.25328042

**Authors:** Gustavo de Araújo Silva, Bruno Wilnes, Beatriz Castello-Branco, José Luiz Padilha da Silva, Marina Pereira Mayrink, Anna Terra França, Marcos Roberto Queiroz França, Reynaldo Castro de Miranda, Maria Carmo Pereira Nunes, Andre Assis Lopes do Carmo

## Abstract

**Background:** Chagas cardiomyopathy (ChC) is a highly arrhythmogenic condition with frequent implantable cardioverter-defibrillator (ICD) therapies and poor clinical outcomes. While catheter ablation is a promising strategy for ventricular tachycardia (VT) management, its effectiveness versus medical therapy in this population remains unclear. This study aimed to evaluate the impact of VT ablation versus medical treatment on VT recurrence and all-cause mortality in patients with ChC and ICDs for secondary prevention, and to compare outcomes between functional and conventional ablation strategies.

**Methods:** In this prospective cohort study, 127 patients with ChC and ICDs were followed for up to two years. Patients were divided into groups according to the treatment received: catheter ablation (n=67) or medical therapy (n=60). The primary outcome was VT recurrence, analyzed with death as a competing risk. Multivariable competing risk and Cox regression models were used.

**Results:** During follow-up, 69 patients had VT recurrence and 40 died. At 24 months, arrhythmia-free survival was 76.2% in the functional ablation group, versus 34.7% and 32.3% in the conventional ablation and medical groups, respectively. Functional ablation was associated with lower risk of VT recurrence (HR: 0.332; 95% CI: 0.121–0.901; p=0.047), and remained protective after adjustment (HR: 0.236; 95% CI: 0.079–0.709; p=0.010). VT recurrence within 30 days post-ablation predicted higher mortality (adjusted HR: 3.247; 95% CI: 1.216–8.665).

**Conclusions:** Catheter ablation, particularly with functional mapping, significantly reduced VT recurrence in ChC patients with ICDs. Early VT recurrence post-ablation independently predicted mortality, underscoring its prognostic importance.

**Clinical Perspective:** *What Is Known:* Patients with Chagas cardiomyopathy experience a high burden of ventricular tachycardia (VT) and frequently require implantable cardioverter-defibrillators (ICDs) for secondary prevention. Catheter ablation is a recognized strategy to reduce VT burden in structural heart disease, but its efficacy in Chagas cardiomyopathy has not been clearly established.

*What The Study Adds:* Adjusted comparisons demonstrated improved survival free of VT recurrence in patients undergoing catheter ablation, with the functional mapping strategy performing better than voltage-based ablation and medical therapy. VT recurrence within 30 days post-ablation was independently associated with increased mortality, highlighting its value as a prognostic marker. These findings support early referral and the use of functional ablation strategies in this high-risk population to improve arrhythmia control and clinical outcomes.

## INTRODUCTION

Chagas disease remains a significant public health challenge, particularly in Latin America, affecting approximately 6 million people globally.^1^ Its more severe manifestation, dilated Chagas cardiomyopathy (ChC), is an infectious cardiomyopathy characterized by a pronounced inflammatory and fibrotic process induced by *T. cruzi* infection, leading to ventricular arrythmias. Ventricular tachycardia ( VT) is highly prevalent in ChC and represents a major cause of mortality, often requiring the use of implantable cardioverter-defibrillators (ICDs) to prevent sudden death. Patients with ChC consistently experience significantly higher rates of appropriate ICD interventions compared to patients with other forms of cardiomyopathy, which can exacerbate ventricular dysfunction and further increase mortality risk. ^2–4^

Effective VT ablation follows a sequence of essential steps, each contributing to a successful outcome. Moreover, recent advancements have introduced innovative techniques that focus on the functional properties of the ventricular substrate, yielding promising results. These new strategies precisely target arrhythmogenic substrates, even in areas that appear to have normal voltage, thereby enhancing treatment efficacy. In our previous study, we demonstrated that functional VT ablation was associated with lower recurrence rates compared to a strategy based on voltage map for identification of arrhythmogenic substrates. ^5^

In addition to ablation, amiodarone is commonly used as part of medical therapy to manage VT in patients with ChC and an implanted ICD. While some studies indicate that amiodarone may reduce VT recurrence and improve outcomes in this population.^6,7^ others have reported an increased risk of mortality associated with its use.^8,9^ Although ICDs play a crucial role in the secondary prevention of sudden cardiac death in ChC, evidence suggests that this intervention does not reduce all-cause mortality in either primary or secondary prevention settings. A recent randomized trial demonstrated that ICDs for primary prevention failed to reduce the risk of all-cause mortality.^10^ These findings highlight the need to focus on strategies that reduce the burden of ventricular arrhythmias and minimize ICD therapies to improve patient outcomes. In this context, ablation therapy emerges as an effective approach for managing ventricular arrhythmias in patients with ICDs, as it significantly reduces arrhythmia recurrence and decreases the rate of ICD interventions. However, its impact on clinical outcomes remains to be fully established and requires further investigation.

Therefore, a rigorous evaluation of VT ablation efficacy compared to amiodarone therapy is essential, especially considering that ablation is not readily accessible for patients with ChC in endemic areas. Notably, there is a lack of comparative studies between ablation and amiodarone therapy in patients with ICDs, and addressing this gap is crucial for optimizing their management.

This study was designed to: 1) assess the impact of VT ablation therapy on VT recurrence and all-cause mortality in patients with ChC who have undergone ICD implantation for secondary prevention of sudden death; 2) compare the outcomes of two distinct ablation therapy approaches; 3) evaluate the prognostic value of 30-day VT recurrence on overall mortality in the subgroup of patients undergoing ablation therapy. Given that death represents a competing event that inherently prevents VT recurrence, this study incorporates competing risks analysis to ensure accurate evaluation of this outcome.

## METHODS

### Study design and population

This study prospectively enrolled a cohort of patients with ChC referred to our institution, a specialized referral center, between 2017 and 2022. Chagas disease was diagnosed based on the presence of at least two positive serological tests for *T. cruzi.* Cardiomyopathy was defined by left ventricular enlargement with systolic dysfunction, typically associated with major electrocardiographic abnormalities.^11^ Patients with ventricular dysfunction caused by other cardiac conditions, including ischemic or idiopathic dilated cardiomyopathy, were excluded.

Eligible participants included patients with ChC who had undergone ICD implantation for secondary prophylaxis and experienced ventricular arrhythmias post-implantation. Among 275 patients with ChC, 165 met the criteria for ICD implantation due to ventricular arrhythmias. Of these, 25 patients were excluded (Figure 1), leaving 127 participants for enrollment. These patients were subsequently divided into two groups based on the treatment approach: Group 1 (cases) received catheter ablation therapy, while Group 2 (controls) were managed with medical treatment using amiodarone. Patients in the ablation group were further subdivided into those undergoing conventional or functional ablation.

**Figure 1.**
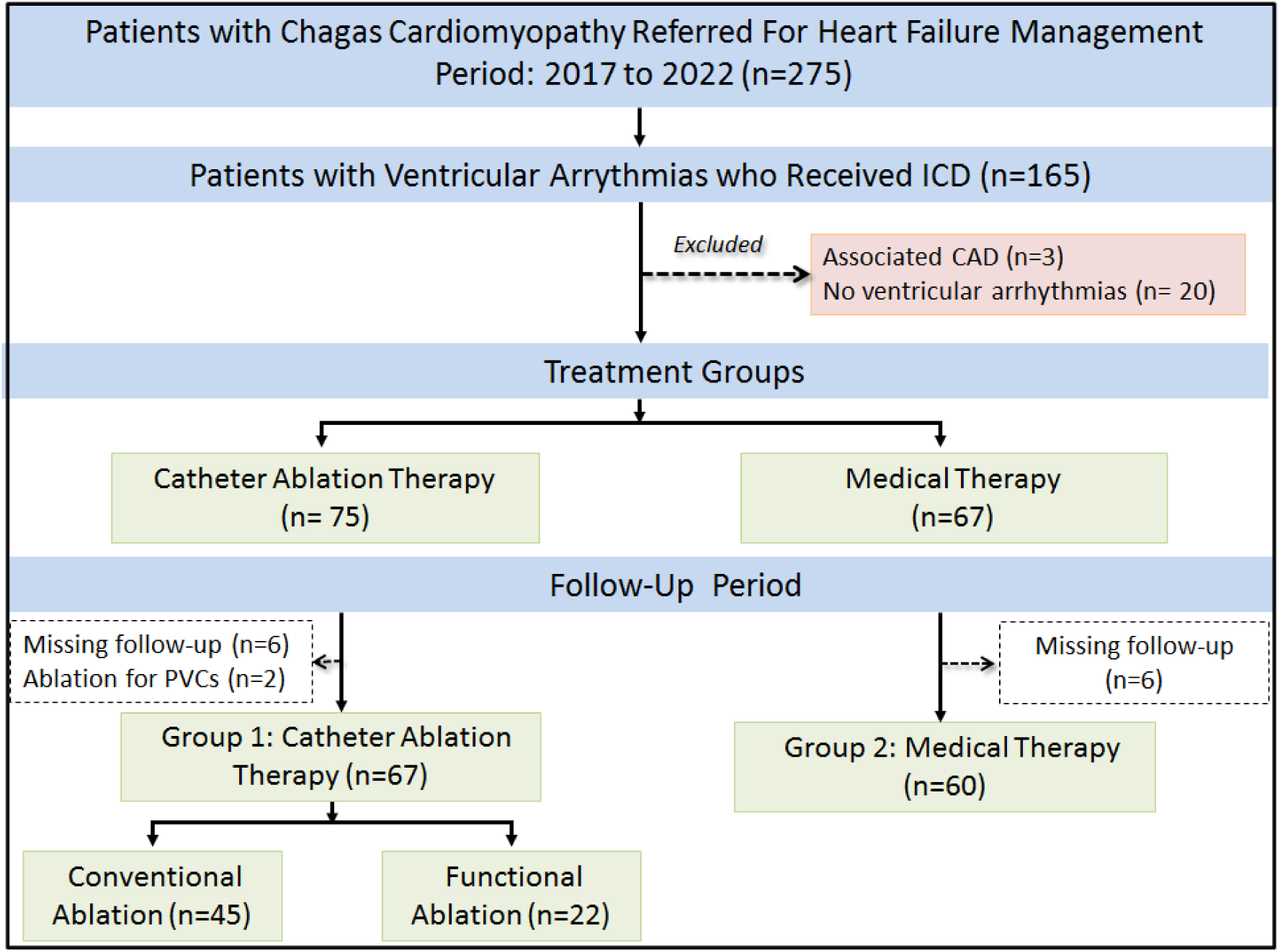
Flowchart illustrating the stratification of patients with Chagas cardiomyopathy (n=275) during the study period. Abbreviations: CAD = coronary artery disease; ICD = implantable cardioverter-defibrillator; VT = ventricular tachycardia; PVC = premature ventricular complex.

### Outcome Definition

Participants were followed for up to 2 years, starting from the date of the sustained VT episode in the medical treatment (control group) or the date of the ablation procedure (case group).

A sustained VT was defined by meeting at least one of the following criteria:

- Ventricular tachycardia or ventricular fibrillation treated by the ICD with antitachycardia pacing (ATP) or shock, including electrical storm (ES), which is defined as three or more sustained VT episodes within a 24-hour period.
- Sustained VT with a heart rate below the ICD detection threshold.

The primary outcome was defined as the recurrence of VT over a two-year follow-up period, with death treated as a competing risk. Secondary outcomes comprised a composite of all-cause mortality and recurrence of VT.

Patients were followed up on an outpatient basis with regular clinical visits and cardiac device evaluations conducted at least every 6 months. ICD programming was based on institutional protocols, and arrhythmic events were systematically recorded in an electronic database. Mortality data was obtained by reviewing medical records, direct contact with family members, or death certificates.

Data collection was conducted utilizing the RedCap platform for longitudinal follow-up and information entry after catheter ablation. Data analysis was also performed using both physical and electronic medical records to ensure comprehensive evaluation. All patients gave written informed consent, and the study protocol was approved by the institutional ethics committee (0682.0.203.000-11)

### Mapping protocol and ablation techniques

All procedures were conducted under general anesthesia. Femoral venous access was established to position catheters in both the right ventricle (RV) and the coronary venous sinus. Right ventricular programmed electrical stimulation (PES) was used to assess VT morphology and evaluate hemodynamic tolerability.^12^ The decision to use intra-aortic balloon pumps (IABP) was guided by the PAINESD risk score, and IABP-assisted procedures were recommended for scores ≥15 or at the operator’s discretion.^13,14^

Our initial approach primarily involved epicardial access using a micropuncture needle, except in patients with a high likelihood of significant adhesions. For left ventricular endocardial access, retrograde arterial and/or transseptal approaches were employed.^15^ Endocardial RV mapping was performed when VT morphology suggested an RV origin or when epicardial mapping revealed scarring in this chamber. Prior to left ventricular endocardial access, systemic anticoagulation with intravenous heparin was administered.^16^

Electroanatomic mapping was performed in all patients. For voltage-based substrate mapping, systems such as EnSite Precision (2-2-2 Livewire or Advisor HD Grid Mapping Catheter Sensor, Abbott Park, IL) or CARTO (Thermocool, Biosense Webster, Diamond Bar, CA) were utilized. EnSite Precision (2-2-2 Livewire or Advisor HD Grid Mapping Catheter Sensor, Abbott Park, IL) was employed for functional mapping due to its capability to record electrograms that allow concurrent construction of various functional maps during ablation procedures.

During epicardial or endocardial left ventricular mapping, RV pacing was the method of choice. Mapping and ablation during VT were initially deferred. Regions containing His electrograms and phrenic nerve locations were identified and marked with distinct color-coded tags.

### Voltage-based and functional substrate mapping in VT ablation

Voltage-based substrate mapping was used to identify ventricular scar tissue during intrinsic rhythm. Bipolar voltage thresholds were used to differentiate normal myocardium (>1.5 mV in the left and right ventricular endocardium; >1.0 mV on the epicardium) from dense scar (<0.5 mV in the left ventricle). Unipolar voltage thresholds (<8.27 mV for left ventricular endocardium,^17^ with lower cutoffs for septal and periaortic regions) further delineated abnormal substrates, including right ventricular epicardial abnormalities (<3.9 mV).^18–20^ Fractionated and late potentials (LPs) were marked to guide ablation. The objective was complete elimination or significant modification of abnormal electrograms, verified post-ablation via programmed electrical stimulation (PES).

Functional mapping was employed to complement voltage-based techniques by assessing dynamic electrophysiological behavior. Single extra-stimulus delivered from the right ventricle (20 ms above the effective refractory period) enabled annotation of late local activation times, with decremental or blocked electrograms identified irrespective of voltage. Isochronal late activation mapping (ILAM),^21,22^ facilitated by advanced mapping tools (e.g., TurboMap), was used to highlight regions of color crowding and delayed activation, critical targets for radiofrequency (RF) ablation. This strategy prioritized modification of epicardial and endocardial LPs to achieve non-inducibility of VT, confirmed through remap of the targeted area and PES.

For persistent inducible VT, activation mapping during tachycardia was utilized to identify protected isthmuses for targeted ablation, contingent on hemodynamic stability. Intrapericardial corticosteroids were administered post-procedure to mitigate pericarditis,^23,24^ and all complications were meticulously recorded to ensure comprehensive procedural evaluation.

### Statistical analysis

Continuous variables were reported as median and interquartile ranges and categorical variables were presented as frequencies and percentage. Baseline characteristics of patients, stratified by treatment group, were compared using the chi-squared test, unpaired Student’s t-test, Mann-Whitney test, depending on the distribution of the variables.

A univariable analysis was performed to assess the impact of ablation therapy on outcomes using the Fine-Gray competing-risks regression model, in which VT recurrence was the primary event and death was treated as a competing risk.

The model was further adjusted for predetermined variables included in the PAINESD score, which incorporated age, NYHA functional class, LV ejection fraction, and the presence of electrical storm. These variables were included in the model as categorical parameters based on the score, including age >60 years, pre-ablation LV ejection fraction <25%, pre-ablation NYHA functional class III or IV and pre-ablation electrical storm. Subsequently, the ablation group was subdivided to analyze the impact of each specific approach on the primary outcome.

Cumulative incidence function (CIF) curves were generated to graphically analyze each variable included in the multivariable model. To better illustrate the pooled results, CIFs were created for each treatment modality, incorporating findings from both univariable and multivariable analyses. The Gray’s test was used to compare cumulative incidences.

In the second analysis, a Cox proportional hazards model was employed to evaluate the prognostic value of 30-day post-ablation VT recurrence on mortality risk. Similar to the Fine-Gray model, the multivariable analysis was adjusted for the predictors included in the PAINESD Score. Additionally, we conducted tests for proportional hazards, interaction assumptions, and multicollinearity among variables, and found no violations. For this analysis, we performed Kaplan-Meier curves and used the log-rank test to compare survival outcomes. A value of p <0.05 was considered statistically significant. All analyses were performed in SPSS version 24 (SPSS Inc., Chicago, Illinois) and in R (R Core Team, 2023), version 4.5.0, using the packages tidyverse, survminer, and cmprsk.

## RESULTS

The baseline characteristics of the patients, categorized into the two groups based on the therapy received, are shown in Table 1. The proportion of females was higher in the ablation group compared to the control group (49% vs. 27%, respectively). There were no significant differences between the groups in terms of age, the severity of left ventricular (LV) systolic dysfunction, medications or pre-existing comorbidities. All patients were receiving optimized heart failure treatment. However, the ablation group exhibited more advanced heart failure, with 37% of patients classified as NYHA functional class III or IV, indicating severe functional limitation despite treatment.

**Table 1:** Baseline characteristics of the study population stratified by treatment group.

| Variables* | Catheter Ablation<br>(n = 67) | Medical Therapy<br>(n = 60) | P<br>value |
| --- | --- | --- | --- |
| Age (years) | 63.0 (55.0-72.0) | 63.5 (54.3-73.0) | 0.987 |
| Male sex | 34 (50.7) | 44 (73.3) | 0.009 |
| NYHA class III/IV | 25 (37.3) | 9 (15.0) | 0.005 |
| LV ejection fraction (%) | 37.0 (30.0-46.0) | 39.0 (32.0-49.0) | 0.214 |
| Comorbidities |  |  |  |
| Hypertension | 16 (23.9) | 22 (36.7) | 0.116 |
| Diabetes | 4 (6.0) | 7 (11.7) | 0.347 |
| eGFR <60 mL/min | 16 (23.9) | 10 (16.7) | 0.314 |
| Atrial fibrillation | 13 (19.4) | 16 (26.7) | 0.330 |
| Thyroidopathy | 23 (34.3) | 19 (31.7) | 0.750 |
| COPD | 4 (6.0) | 4 (6.7) | 1.000 |
| Stroke | 7 (10.4) | 6 (10.0) | 1.000 |
| Medications |  |  |  |
| Beta-blockers | 65 (97.0) | 57 (95.0) | 0.667 |
| ACEI or ARB | 51 (76.1) | 46 (76.7) | 0.942 |
| Propafenone | 1 (1.5) | 0 (0) | 1.000 |
| Espironolactone | 34 (50.7) | 26 (43.3) | 0.403 |
| Anticoagulation | 7 (10.4) | 24 (40.0) | <0.001 |
| Diuretic | 32 (47.8) | 33 (55.0) | 0.415 |
| Amiodarone use | 60 (89.6) | 49 (83.1) | 0.286 |
| Amiodarone dose (mg) | 400 (400-800) | 200 (100-200) | <0.001 |
| Previous ventricular arrhythmia |  |  |  |
| Incessant ventricular tachycardia | 11 (16.4) | 1 (1.8) | 0.006 |
| Ventricular tachycardia below the detection zone | 21 (31.3) | 9 (15.3) | 0.034 |
| Ventricular tachycardia storm | 58 (86.6) | 10 (16.7) | <0.001 |

\*Data are expressed as the median (inter-quartile range) or number (percentage) of patients
Abbreviations: ACEI: Angiotensin-converting–enzyme inhibitor; ARB: Angiotensin-receptor blocker; COPD: Chronic obstructive pulmonary disease; eGFR: Estimated glomerular filtration rate; LV: Left ventricle.

Although amiodarone use was comparable between the groups, patients in the ablation group received higher baseline doses of the medication (400 mg daily, ranging from 400 to 800 mg). Only one patient in the ablation group was taking propafenone.

As anticipated, the proportion of patients with ventricular arrhythmias at baseline was higher in the ablation group, with incessant VT and VT below the detection zone observed in 16% and 31% of patients, respectively. Notably, electrical storms occurred in 87% of the ablation group compared to 17% in the medical treatment group. A subanalysis comparing patient characteristics between the conventional and functional ablation subgroups revealed no significant differences.

### Clinical outcomes

During a 2-year follow-up period (IQR 2.2 - 24 months), 69 patients experienced VT recurrence as their first event. Among them, 26 subsequently died following VT recurrence. Additionally, 14 patients who did not experience VT recurrence died, with death considered a competing risk preventing the occurrence of VT recurrence. In total, 83 composite outcomes were recorded. In the competing risk analysis, comparing medical therapy with catheter ablation, the incidence of VT recurrence was similar between groups, with a hazard ratio (HR) of 0.695 (95% CI: 0.432–1.120) (Figure 2). Arrhythmia-free survival rates at 12 months were 52.0% [95% CI: 38.2%–64.1%] in the general ablation group and 52.5% [95% CI: 39.1%–64.3%] in the clinical treatment group. At 24 months, the rates were 48.0% [95% CI: 34.3%–60.4%] and 32.3% [95% CI: 20.6%–44.6%], respectively. However, in patients with electrical storm, ablation therapy was strongly associated with a reduced risk of VT recurrence, with an HR of 0.453 (95% CI: 0.224–0.923) (Figure 3).

**Figure 2.**
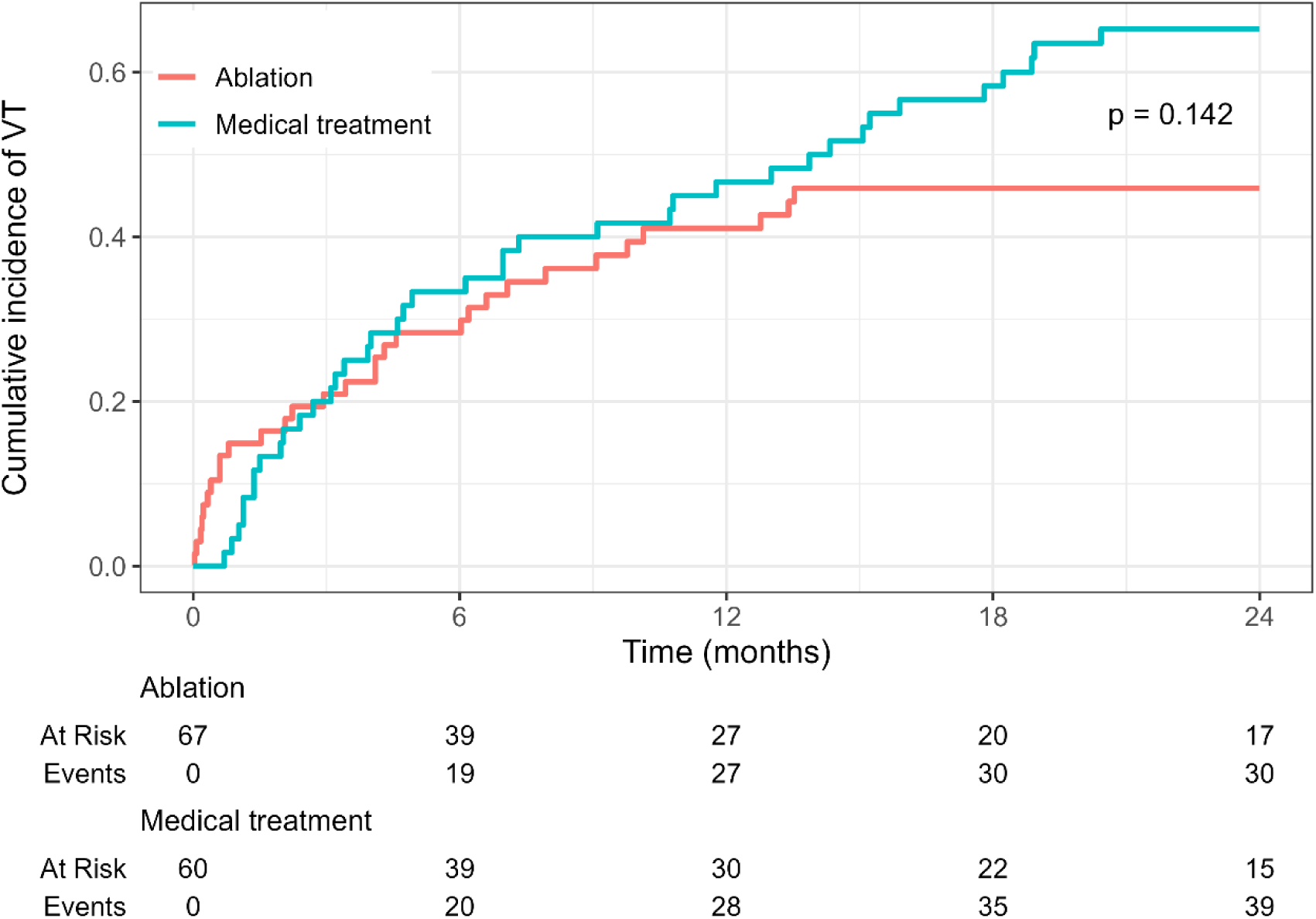
Cumulative incidence of ventricular tachycardia recurrence over 24 months in patients with Chagas cardiomyopathy undergoing ablation compared with medical therapy. Death treated as a competing risk.

**Figure 3.**
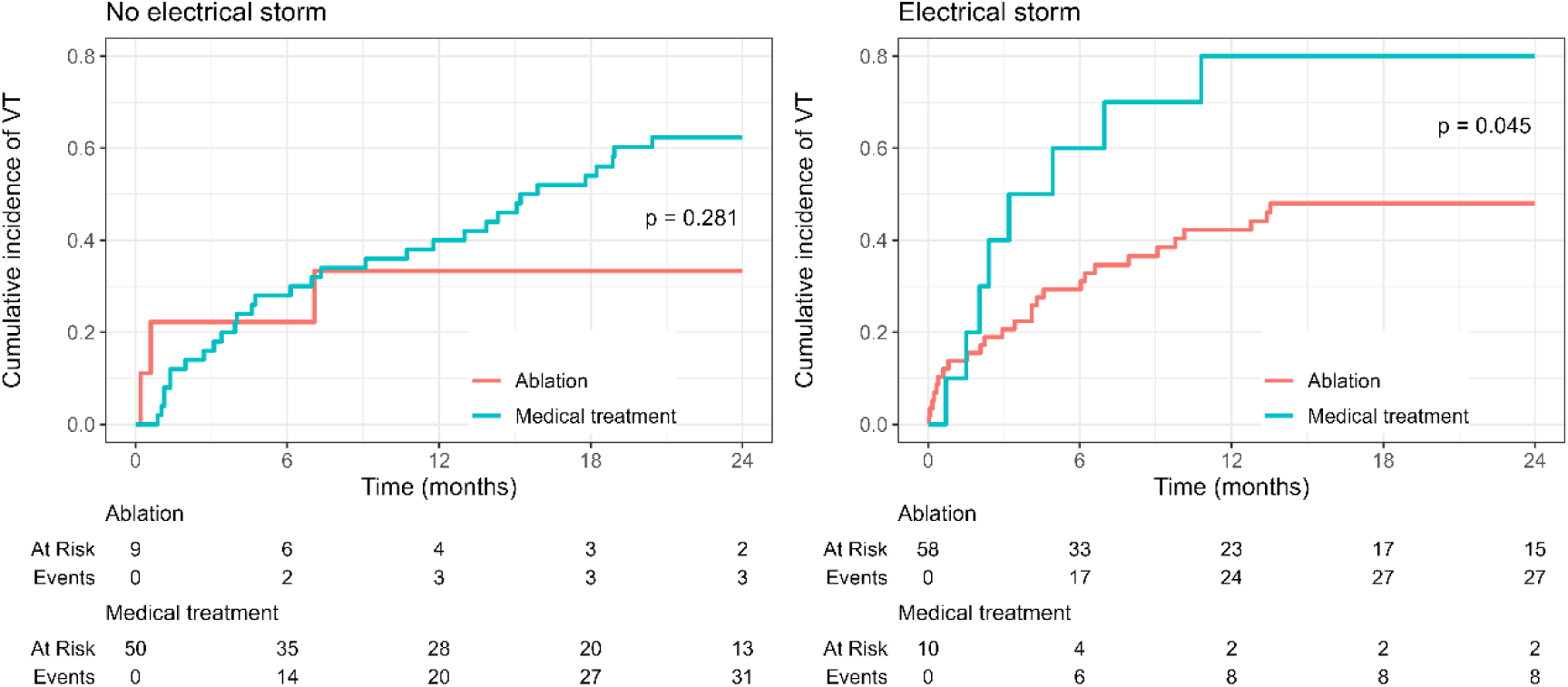
Cumulative incidence of ventricular tachycardia recurrence stratified by the presence of electrical storm, with death treated as a competing risk. Ablation was associated with a significantly lower risk of ventricular tachycardia recurrence in patients presenting with electrical storm (HR: 0.453; 95% CI: 0.224–0.923, right panel), whereas no significant benefit was observed in those without electrical storm (left panel).

Regarding the competitive risk analysis, when comparing medical therapy with conventional or functional ablation, a lower recurrence of ventricular arrhythmia was observed in patients who underwent functional ablation (HR of 0.332, 95% CI 0.121-0.901, p=0.047) (Figure 4). Arrhythmia-free survival rates at 12 months were 76.2% [95% CI: 51.8%–89.4%] in the functional ablation group, 40.5% [95% CI: 24.8%–55.6%] in the conventional ablation group, and 52.5% [95% CI: 39.1%–64.3%] in the clinical treatment group. At 24 months, the rates were 76.2% [95% CI: 51.8%–89.4%], 34.7% [95% CI: 19.9%–49.9%], and 32.3% [95% CI: 20.6%–44.6%], respectively. When comparing the functional ablation, conventional ablation, and medical treatment groups, the combined outcome of freedom from death or ventricular arrhythmia recurrence were 68.2% vs. 26.7% vs. 28.0% (p=0.030), demonstrating a statistically significant difference, with the functional ablation group maintaining a higher proportion of patients free from recurrence or death during the 24-month follow-up.

**Figure 4.**
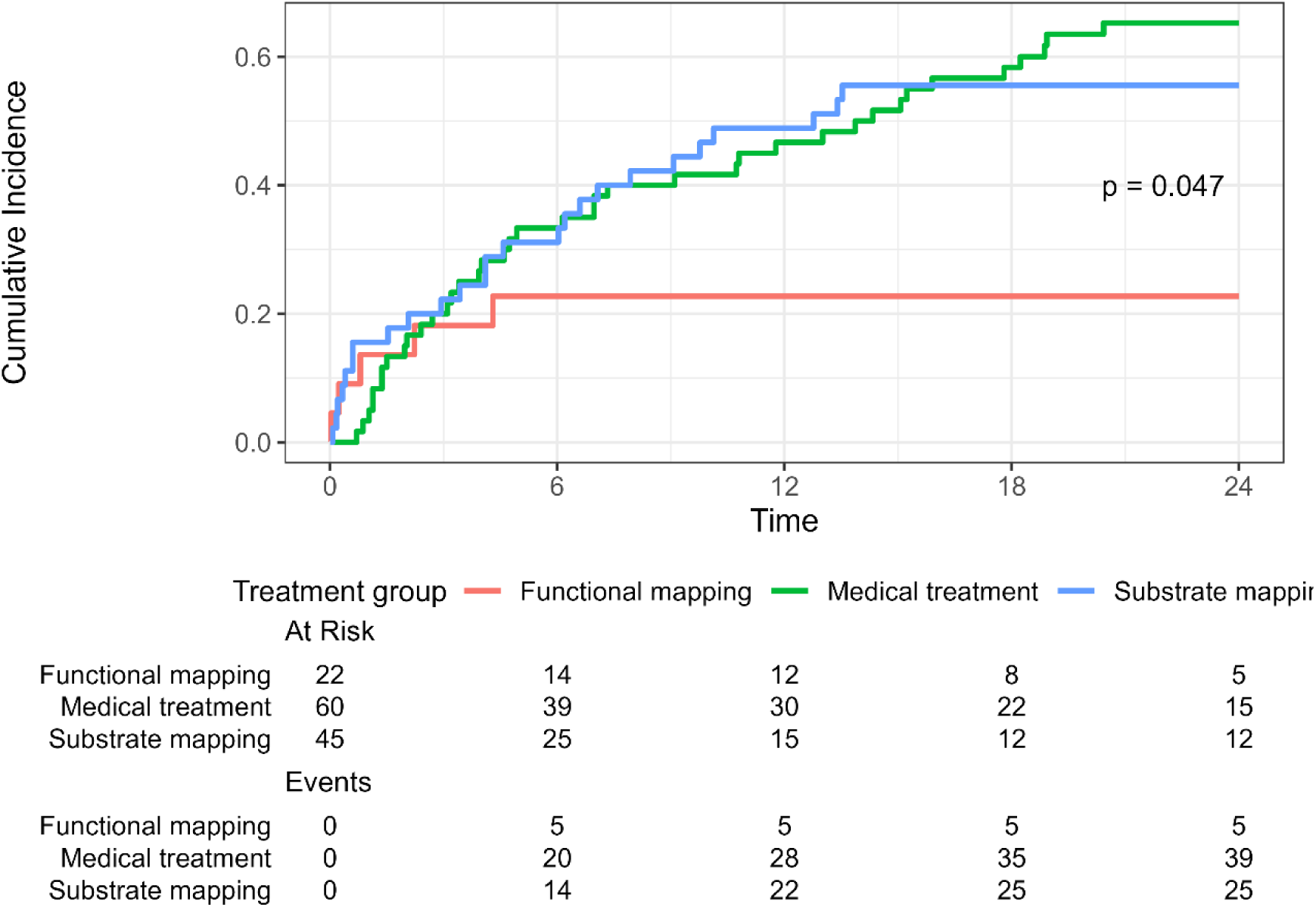
Cumulative incidence of ventricular tachycardia recurrence across treatment strategies: functional mapping, substrate mapping, and medical therapy. Death was treated as a competing risk. Functional mapping was associated with significantly lower ventricular tachycardia recurrence (HR of 0.332, 95% CI 0.121-0.901)

Moreover, in the multivariable model adjusting initially by electrical storm, the impact of ablation was evident as a protective factor of VT recurrence (Figure 5). Subsequently, after adjustment for the variables included in the PAINSED score, catheter ablation remained as a predictor of reduced VT recurrence, with an HR of 0.441 (95% CI: 0.217–0.926) (Figure 5A). The approach using functional mapping, when analyzed through this multivariable model, reinforced the technique as a protective factor against VT recurrence HR 0.236 (95% CI 0.079-0.709, P=0.010) (Figure 5B), this indicates a 76.4% reduction in arrhythmia recurrence when functional catheter ablation was performed. All-cause mortality was similar between the groups, with event-free survival of 62% in the case group and 72% in the control group (p=0.100).

**Figure 5.**
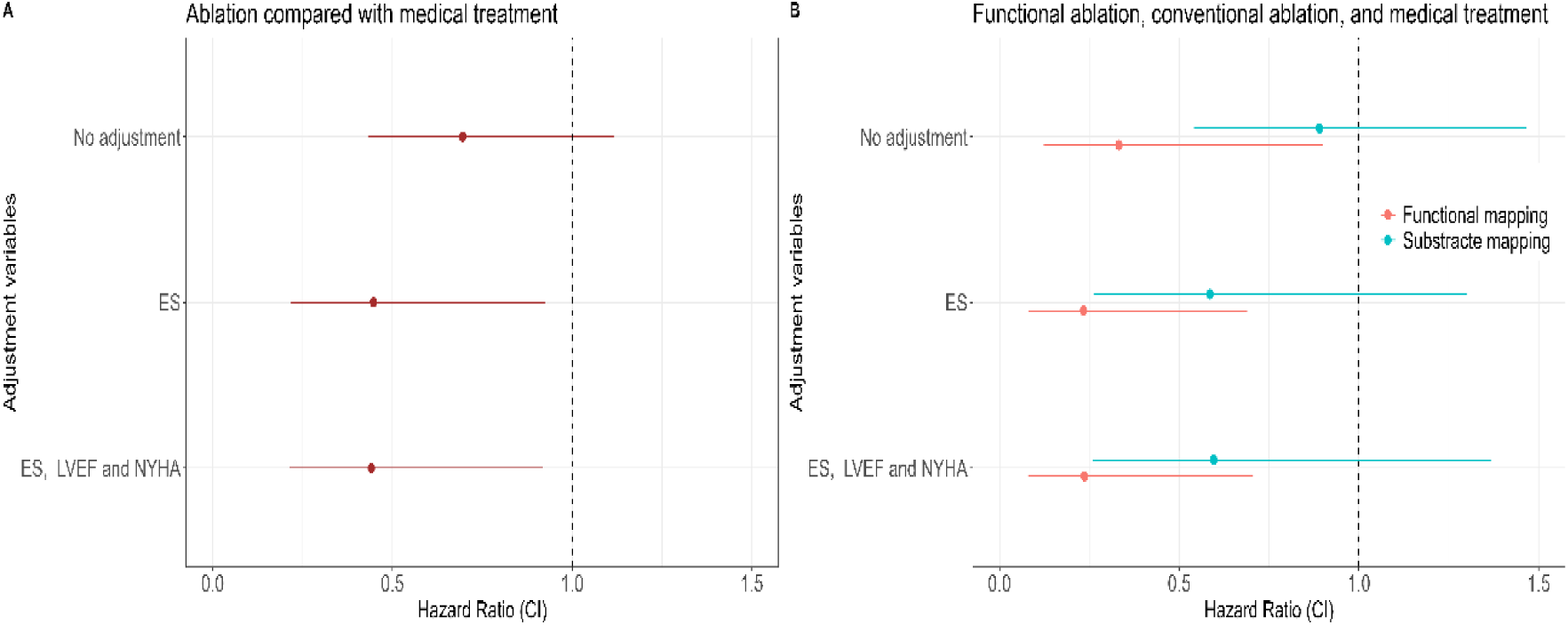
**(A-B)**. Multivariable hazard models evaluating the impact of ablation on VT recurrence, using the Fine-Gray model with death as a competing risk. **A:** Comparison of catheter ablation versus medical therapy. In the unadjusted analysis, VT recurrence rates were similar between groups. After adjustment for electrical storm at presentation, ablation was associated with a reduced risk of VT recurrence. This association remained significant after further adjustment for LVEF and NYHA functional class. **B:** Comparison of ablation strategies. Functional mapping was consistently associated with a lower risk of VT recurrence compared to both conventional (substrate-based) ablation and medical therapy, in both unadjusted and adjusted models. Abbreviations: CI = confidence interval, ES = electrical storm; LVEF = left ventricular ejection fraction; NYHA = New York Heart Association functional class, VT = ventricular tacxhycardia.

Further analysis comparing functional mapping, voltage-based ablation, and medical therapy revealed that the functional catheter mapping approach was associated with better outcomes. Specifically, it was linked to a lower risk of VT recurrence in both unadjusted and adjusted models, accounting for variables included in the PAINSED score (HR 0.332, 95% CI 0.121-0.901).

A subanalysis was conducted to assess the impact of VT recurrence within 30 days post-catheter ablation on mortality rates, observed in 15% of patients. In the univariable Cox proportional hazards regression model, early arrhythmia recurrence was associated with a significantly increased risk of mortality (HR: 5.570;95% CI 2.230–13.915) (Figure 6). After adjusting for age, sex, and key prognostic variables, including NYHA class, LV ejection fraction, and the presence of an electrical storm, early arrhythmia recurrence remained a strong independent predictor of mortality (HR: 3.247; 95% CI 1.216-8.665) (Table 2). Additionally, LV ejection fraction was a powerful predictor of mortality in this population (HR 0.901: 95% CI 0.853-0.952).

**Figure 6.**
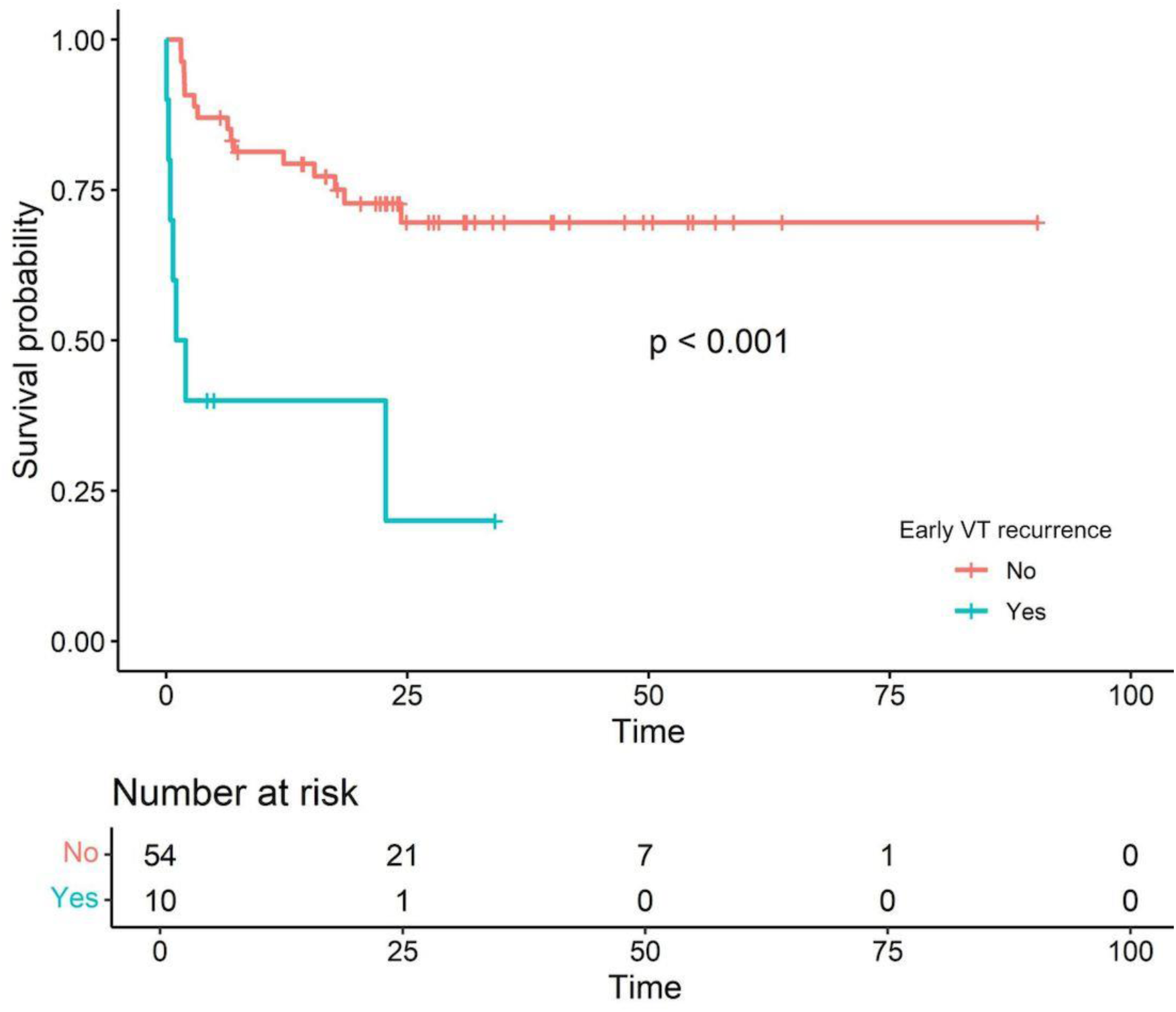
Kaplan–Meier survival curves comparing patients with and without early VT recurrence (≤30 days post-ablation). Early VT recurrence was associated with significantly higher mortality rate (HR of 5.570;95% CI 2.230– 13.915). Abbreviations: HR = hazard ratio, VT = ventricular tachycardia.

**Table 2:** Predictors of Mortality in Patients Undergoing Ablation Therapy.

| Variables | Unadjusted |  | Multivariable adjustment for age, sex, NYHA class, LVEF, electrical storm, and 30-day VT recurrence |  |
| --- | --- | --- | --- | --- |
|  | Hazard ratio (95% CI) | P value | Hazard ratio (95% CI) | P value |
| Age (Years) | 1.003 (0.976-1.031) | 0.811 | ... | ... |
| Male | 0.977 (0.522-1.830) | 0.942 | ... | ... |
| NYHA class III/IV | 3.814 (2.063-7.053) | <0.001 | ... | ... |
| LVEF (%) | 0.949 (0.920-0.979) | 0.001 | 0.901 (0.853-0.952) | <0.001 |
| Electrical storm* | 2.044 (1.071-3.901) | 0.030 | ... | ... |
| 30-day VT recurrence | 5.570 (2.230-13.915) | <0.001 | 3.247 (1.216-8.665) | 0.019 |

\*Electrical storm pre ablation
Abbreviations:
CI = confidence interval; LVEF = left ventricular ejection fraction; VT = ventricular tachycardia

## DISCUSSION

In this study involving a well-established cohort of patients with ChC requiring ICDs, several key findings were identified. First, the recurrence rate of VT was notably high, with over half of the patients experiencing VT recurrence at 2-year follow-up, underscoring the highly arrhythmogenic nature of ChC. Second, VT ablation therapy was associated with a significantly lower incidence of VT recurrence and mortality, after adjusting for the presence of electrical storms and other prognostic variables, using competing risks analysis to ensure accurate estimates. Third, a functional ablation approach was independently associated with a reduced risk of VT recurrence, regardless of electrical storm occurrence or heart failure (HF) severity. Finally, in patients who underwent ablation, VT recurrence within 30 days post-procedure emerged as a strong independent predictor of mortality. Taking together, these findings provide important insights into optimizing treatment strategies for VT in patients with ChC and highlight the potential benefits of early and targeted ablation therapy.

This study addresses a critical gap in the literature by directly comparing the outcomes of catheter ablation versus medical therapy in this high-risk population. Notably, the clinical profile of patients undergoing ablation highlights the barriers to accessing specialized VT ablation centers. Consequently, ablation procedures are frequently performed on patients with more advanced disease stages and poorer prognoses. These patients frequently present with severe HF, as indicated by a higher prevalence of NYHA class IV, and exhibit a greater incidence of life-threatening arrhythmic events with multiple ICD shocks.

The management of ventricular arrhythmias in patients with ChC remains a significant clinical challenge. Although ICDs are often lifesaving, they do not prevent ventricular arrhythmias, and recurrent appropriate shocks, while effective in terminating arrhythmias, can significantly impact patients’ quality of life and have also been associated with increased mortality.^25–28^ To reduce arrhythmic burden, amiodarone is commonly used; however, its variable effectiveness and potential for serious adverse effects limit its long-term utility in clinical practice.^29–31^

In this context, catheter ablation has emerged as a promising strategy to reduce VT recurrence in patients with structural heart disease. Despite its potential benefits, there is still a lack of robust data on the impact of VT ablation on recurrence rates and overall survival in patients with ChC, underscoring the need for further research in this population.

ChC is an inherently arrhythmogenic condition associated with a worse prognosis than other forms of HF. The poor outcomes observed in ChC-related HF are largely driven by the high prevalence of complex ventricular arrhythmias, which remain the leading cause of sudden cardiac death in this population.^32,33^ Strategies to prevent sudden death in ChC have primarily relied on the empirical use of amiodarone, ICDs, or a combination of both.

Although ICDs effectively reduce the risk of sudden death, their impact on overall survival remains uncertain. A recent randomized clinical trial involving 323 high-risk patients (157 in the ICD group and 166 in the amiodarone group) demonstrated no significant reduction in all-cause mortality with ICD therapy.^10^ Similarly, a meta-analysis conducted by our group, which focused on patients with ChC who received an ICD exclusively for secondary prevention, found no significant reduction in all-cause mortality compared to treatment with amiodarone.^34^

These findings highlight a critical limitation: while ICDs effectively prevent sudden cardiac death, they do not reduce mortality from progressive HF, which remains the leading cause of death in this patient population. Our previous studies reinforce this observation, showing that in patients with ChC and severe ventricular dysfunction, HF is the predominant mode of death.^35–37^ A meta-analysis further confirmed that the mode of death in ChC is strongly influenced by the clinical profile of the studied population. In cohorts composed predominantly of asymptomatic, ambulatory patients, those in NYHA class I/II, or individuals with documented ventricular arrhythmias not treated with an ICD, sudden cardiac death rates exceeded those from HF. In contrast, among patients with symptomatic chronic ChC, left ventricular dysfunction or overt HF, and those treated with ICD or cardiac resynchronization therapy (CRT), death from progressive HF was more frequent.^38^These insights underscore the need for a more comprehensive treatment approach that not only targets ventricular arrhythmias but also addresses the underlying myocardial dysfunction driving HF progression in ChC. In this context, catheter ablation may represent a valuable therapeutic option, offering a more effective strategy for managing ventricular arrhythmias and potentially improving overall outcomes.

### VT ablation in nonischemic cardiomyopathy

Recent advancements in ablation techniques, particularly the use of functional mapping, have enhanced our understanding of scar tissue and arrhythmogenic circuits, enabling more precise and effective ablation procedures. A recent study by our group demonstrated that functional mapping-guided ablation, compared to conventional techniques, reduced VT recurrence by 90.3%.^5^ Consequently, this study also examined the technical distinctions between different ablation approaches.

As previously demonstrated by our group, functional mapping techniques have significantly improved VT recurrence outcomes following ablation. In particular, extra-stimuli-assisted functional mapping has shown a substantial reduction in VT recurrence rates compared to static functional mapping techniques in both ischemic and non-ischemic cardiomyopathies.^39^

The use of extra-stimuli enhances the ability to uncover concealed regions of decremental or slow conduction, which are often critical for VT initiation and maintenance. Moreover, extra-stimuli can simulate VT induction, providing valuable insights into conduction patterns around scarred areas. For patients with extensive arrhythmogenic substrates, such as those with ChC, these techniques are especially beneficial, as they allow for a more comprehensive substrate modification. In this study, we observed a two-year VT recurrence rate of 23,8%, highlighting the effectiveness of this approach in a population with severe disease burden.

This study did not find difference in mortality rates between medical treatment and ablation groups, which is a relevant finding considering the presence of baseline characteristics associated with a worse prognosis in the catheter ablation group. The subgroup analysis evaluating the association between ventricular arrhythmia recurrence within the first 30 days after ablation showed that lack of arrhythmia control was associated with increased mortality (HR: 5.570; 95% CI: 2.230–13.915).

The poorer prognosis of patients with ventricular arrhythmia recurrence, combined with the intrinsic characteristics of ChC – such as a higher arrhythmic burden and worse clinical outcomes – highlights the need for optimized treatment strategies. Advances in catheter ablation techniques, particularly functional mapping, offer a promising approach in this context. This study reinforces the safety and efficacy of functional mapping-guided ablation as a valuable therapeutic option for managing ventricular arrhythmia in patients with ChC.

Our study has several limitations. First, there was a discrepancy in the timing of the ablation procedures between the conventional and functional mapping groups. Patients in the conventional ablation group, who underwent voltage-based substrate mapping, received VT ablation earlier than those in the functional mapping group. Additionally, the observational design of the study inherently limits its ability to establish causality and introduces potential biases that must be carefully considered.

This study also has several notable strengths. All procedures in both groups were performed collaboratively by two experienced cardiac electrophysiologists, minimizing inter-procedural variability and enhancing data reproducibility. Furthermore, ChC is a highly arrhythmogenic condition, and our cohort included patients with a markedly severe baseline clinical profile. Consequently, our analysis may underestimate the true benefits of VT ablation, which could be even more pronounced in a population with a less severe presentation of ChC. Lastly, the use of rigorous statistical methodology allowed for the development of well-founded hypotheses regarding the advantages of functional mapping techniques for improving outcomes after VT ablation.

## CONCLUSION

This study demonstrates that patients with Chagas cardiomyopathy requiring ICDs face a high burden of VT recurrence, driven by the disease’s inherently arrhythmogenic substrate. Catheter ablation, particularly with a functional approach, significantly reduced the combined endpoint of VT recurrence and mortality, independent of electrical storms and heart failure severity. Early VT recurrence within 30 days post-ablation emerged as a strong predictor of mortality, emphasizing the importance of close post-procedural monitoring. Moreover, patients undergoing ablation often presented with more advanced heart failure and poorer prognoses, indicating the need for earlier intervention and improved access to specialized care to optimize outcomes in this high-risk population.

## Non-standard Abbreviations and Acronyms

ChC: Chagas cardiomyopathy
VT: Ventricular tachycardia
ICD: Implantable cardioverter-defibrillator
PES: Programmed electrical stimulation
ILAM: Isochronal Late Activation Mapping
LP: Late potential
DZ: Deceleration zone
CIF: Cumulative incidence function
PVC: Premature ventricular complex
RF: Radiofrequency

## Data Availability

The specific algorithms, codes, and software used in data analysis and interpretation are available from the corresponding author upon reasonable request. The authors are committed to transparency and reproducibility, providing necessary information for verification and further research.

## REFERENCES

1. Oliveira GMM de, Brant LCC, Polanczyk CA, et al. Estatística Cardiovascular – Brasil 2020. Arq Bras Cardiol. 2020;115(3):308–439. doi:10.36660/abc.20200812

2. Barbosa MPT, da Costa Rocha MO, de Oliveira AB, Lombardi F, Ribeiro ALP. Efficacy and safety of implantable cardioverter-defibrillators in patients with Chagas disease. EP Europace. 2013;15(7):957–962. doi:10.1093/europace/eut011

3. FILHO MM, DE SIQUEIRA SF, MOREIRA H, et al. Probability of Occurrence of Life-Threatening Ventricular Arrhythmias in Chagas’ Disease versus Non-Chagas’ Disease. Pacing and Clinical Electrophysiology. 2000;23(11P2):1944–1946. doi:10.1111/j.1540-8159.2000.tb07058.x

4. França AT, Martins LNA, de Oliveira DM, et al. Evaluation of patients with implantable cardioverter–defibrillator in a Latin American tertiary center. J Cardiovasc Electrophysiol. 2024;35(4):675–684. doi:10.1111/jce.16201

5. Wilnes B, Castello-Branco B, Silva GA, et al. Enhancing Ventricular Tachycardia Ablation Outcomes. JACC Clin Electrophysiol. 2025;11(1):200–202. doi:10.1016/j.jacep.2024.09.030

6. Morillo CA, Marin-Neto JA, Avezum A, et al. Randomized Trial of Benznidazole for Chronic Chagas’ Cardiomyopathy. New England Journal of Medicine. 2015;373(14):1295–1306. doi:10.1056/NEJMoa1507574

7. Stein C, Migliavaca CB, Colpani V, et al. Amiodarone for arrhythmia in patients with Chagas disease: A systematic review and individual patient data meta-analysis. PLoS Negl Trop Dis. 2018;12(8):e0006742. doi:10.1371/journal.pntd.0006742

8. Bardy GH, Lee KL, Mark DB, et al. Amiodarone or an Implantable Cardioverter–Defibrillator for Congestive Heart Failure. New England Journal of Medicine. 2005;352(3):225–237. doi:10.1056/NEJMoa043399

9. Torp-Pedersen C, Metra M, Spark P, et al. The Safety of Amiodarone in Patients With Heart Failure. J Card Fail. 2007;13(5):340–345. doi:10.1016/j.cardfail.2007.02.009

10. Martinelli-Filho M, Marin-Neto JA, Scanavacca MI, et al. Amiodarone or Implantable Cardioverter-Defibrillator in Chagas Cardiomyopathy. JAMA Cardiol. 2024;9(12):1073. doi:10.1001/jamacardio.2024.3169

11. Bern C. Chagas’ Disease. New England Journal of Medicine. 2015;373(5):456–466. doi:10.1056/NEJMra1410150

12. O’DONNELL D, BOURKE JP, FURNISS SS. Standardized Stimulation Protocol to Predict the Long–Term Success of Radiofrequency Ablation of Postinfarction Ventricular Tachycardia. Pacing and Clinical Electrophysiology. 2003;26(1p2):348–351. doi:10.1046/j.1460-9592.2003.00047.x

13. Santangeli P, Frankel DS, Tung R, et al. Early Mortality After Catheter Ablation of Ventricular Tachycardia in Patients With Structural Heart Disease. J Am Coll Cardiol. 2017;69(17):2105–2115. doi:10.1016/j.jacc.2017.02.044

14. Muser D, Liang JJ, Castro SA, et al. Outcomes with prophylactic use of percutaneous left ventricular assist devices in high-risk patients undergoing catheter ablation of scar-related ventricular tachycardia: A propensity-score matched analysis. Heart Rhythm. 2018;15(10):1500–1506. doi:10.1016/j.hrthm.2018.04.028

15. Lesh MD, Van Hare GF, Scheinman MM, Ports TA, Epstein LA. Comparison of the retrograde and transseptal methods for ablation of left free wall accessory pathways. J Am Coll Cardiol. 1993;22(2):542–549. doi:10.1016/0735-1097(93)90062-6

16. Muser D, Liang JJ, Castro SA, et al. Outcomes with prophylactic use of percutaneous left ventricular assist devices in high-risk patients undergoing catheter ablation of scar-related ventricular tachycardia: A propensity-score matched analysis. Heart Rhythm. 2018;15(10):1500–1506. doi:10.1016/j.hrthm.2018.04.028

17. Hutchinson MD, Gerstenfeld EP, Desjardins B, et al. Endocardial Unipolar Voltage Mapping to Detect Epicardial Ventricular Tachycardia Substrate in Patients With Nonischemic Left Ventricular Cardiomyopathy. Circ Arrhythm Electrophysiol. 2011;4(1):49–55. doi:10.1161/CIRCEP.110.959957

18. Liang JJ, D’Souza BA, Betensky BP, et al. Importance of the Interventricular Septum as Part of the Ventricular Tachycardia Substrate in Nonischemic Cardiomyopathy. JACC Clin Electrophysiol. 2018;4(9):1155–1162. doi:10.1016/j.jacep.2018.04.016

19. Nakajima I, Narui R, Aboud AA, et al. Periaortic Ventricular Tachycardias in Nonischemic Cardiomyopathy. Circ Arrhythm Electrophysiol. 2021;14(2). doi:10.1161/CIRCEP.120.008887

20. Venlet J, Piers SRD, Kapel GFL, et al. Unipolar Endocardial Voltage Mapping in the Right Ventricle. Circ Arrhythm Electrophysiol. 2017;10(8). doi:10.1161/CIRCEP.117.005175

21. Irie T, Yu R, Bradfield JS, et al. Relationship Between Sinus Rhythm Late Activation Zones and Critical Sites for Scar-Related Ventricular Tachycardia. Circ Arrhythm Electrophysiol. 2015;8(2):390–399. doi:10.1161/CIRCEP.114.002637

22. Raiman M, Tung R. Automated isochronal late activation mapping to identify deceleration zones: Rationale and methodology of a practical electroanatomic mapping approach for ventricular tachycardia ablation. Comput Biol Med. 2018;102:336–340. doi:10.1016/j.compbiomed.2018.07.012

23. Dyrda K, Piers SRD, van Huls van Taxis CF, Schalij MJ, Zeppenfeld K. Influence of Steroid Therapy on the Incidence of Pericarditis and Atrial Fibrillation After Percutaneous Epicardial Mapping and Ablation for Ventricular Tachycardia. Circ Arrhythm Electrophysiol. 2014;7(4):671–676. doi:10.1161/CIRCEP.113.001148

24. D’AVILA A, NEUZIL P, THIAGALINGAM A, et al. Experimental Efficacy of Pericardial Instillation of Anti–inflammatory Agents during Percutaneous Epicardial Catheter Ablation to Prevent Postprocedure Pericarditis. J Cardiovasc Electrophysiol. 2007;18(11):1178–1183. doi:10.1111/j.1540-8167.2007.00945.x

25. Knops RE, de Veld JA, Ghani A, et al. Quality of Life in Subcutaneous or Transvenous Implantable Cardioverter-Defibrillator Patients: A Secondary Analysis of the PRAETORIAN Trial. Circ Cardiovasc Qual Outcomes. 2024;17(11):e010822. doi:10.1161/CIRCOUTCOMES.124.010822

26. Januszkiewicz Ł, Barra S, Providencia R, et al. Long-term quality of life and acceptance of implantable cardioverter-defibrillator therapy: results of the European Heart Rhythm Association survey. EP Europace. 2022;24(5):860–867. doi:10.1093/europace/euac011

27. Sears SF, Rosman L, Sasaki S, et al. Defibrillator shocks and their effect on objective and subjective patient outcomes: Results of the PainFree SST clinical trial. Heart Rhythm. 2018;15(5):734–740. doi:10.1016/j.hrthm.2017.12.026

28. Guerra F, Shkoza M, Scappini L, Flori M, Capucci A. Role of electrical storm as a mortality and morbidity risk factor and its clinical predictors: a meta-analysis. Europace. 2014;16(3):347–353. doi:10.1093/europace/eut304

29. Scanavacca MISEALJHBGPF. Terapêutica empírica com amiodarona em portadores de miocardiopatia chagásica crônica e taquicardia ventricular sustentada / Long term results of empiric amiodarone therapy in patients with sustained ventricular tachycardia and chronic chagasic myocarditis. Arq Bras Cardiol. 1990;54(6):367–371.

30. Bardy GH, Lee KL, Mark DB, et al. Amiodarone or an Implantable Cardioverter–Defibrillator for Congestive Heart Failure. New England Journal of Medicine. 2005;352(3):225–237. doi:10.1056/NEJMoa043399

31. Wiedmann F, Ince H, Stellbrink C, et al. Single beta-blocker or combined amiodarone therapy in implantable cardioverter-defibrillator and cardiac resynchronization therapy—defibrillator patients: Insights from the German DEVICE registry. Heart Rhythm. 2023;20(4):501–509. doi:10.1016/j.hrthm.2022.12.009

32. Barbosa MPT, Carmo AAL do, Rocha MO da C, Ribeiro ALP. Ventricular arrhythmias in Chagas disease. Rev Soc Bras Med Trop. 2015;48(1):4–10. doi:10.1590/0037-8682-0003-2014

33. Freitas HFG, Chizzola PR, Paes ÂT, Lima ACP, Mansur AJ. Risk stratification in a Brazilian hospital-based cohort of 1220 outpatients with heart failure: role of Chagas’ heart disease. Int J Cardiol. 2005;102(2):239–247. doi:10.1016/j.ijcard.2004.05.025

34. Carmo AAL, de Sousa MR, Agudelo JF, et al. Implantable cardioverter-defibrillator in Chagas heart disease: A systematic review and meta-analysis of observational studies. Int J Cardiol. 2018;267:88–93. doi:10.1016/j.ijcard.2018.05.091

35. Nunes M do CP, Rocha MOC, Ribeiro ALP, et al. Right ventricular dysfunction is an independent predictor of survival in patients with dilated chronic Chagas’ cardiomyopathy. Int J Cardiol. 2008;127(3):372–379. doi:10.1016/j.ijcard.2007.06.012

36. Nunes MCP, Barbosa MM, Ribeiro ALP, Colosimo EA, Rocha MOC. Left Atrial Volume Provides Independent Prognostic Value in Patients With Chagas Cardiomyopathy. Journal of the American Society of Echocardiography. 2009;22(1):82–88. doi:10.1016/j.echo.2008.11.015

37. Nunes MP, Colosimo EA, Reis RCP, et al. Different prognostic impact of the tissue Doppler-derived E/e′ ratio on mortality in Chagas cardiomyopathy patients with heart failure. The Journal of Heart and Lung Transplantation. 2012;31(6):634–641. doi:10.1016/j.healun.2012.01.865

38. Chadalawada S, Rassi A, Samara O, et al. Mortality risk in chronic Chagas cardiomyopathy: a systematic review and meta–analysis. ESC Heart Fail. 2021;8(6):5466–5481. doi:10.1002/ehf2.13648

39. Wilnes B, Castello-Branco B, Pereira EMM, et al. Extrastimuli-assisted Functional Mapping Improves Ventricular Tachycardia Ablation Outcomes: A Systematic Review, Meta-analysis, and Meta-regression. Heart Rhythm. Published online April 2025. doi:10.1016/j.hrthm.2025.03.2000

